# AI-Powered Exploration of IGF2BP3 as a Prognostic Biomarker in Chronic Myeloid Leukemia Progression and Disease Stratification

**DOI:** 10.1101/2025.01.02.24319743

**Authors:** Pragati Chauhan, Saba Ubaid, Mohammad Kashif, Tanvi Singh, Gaurav Singh, Shailendra Prasad Verma, Ranjana Singh, Rashmi Kushwaha, Vivek Singh

## Abstract

**Background:** Chronic Myeloid Leukemia (CML) progresses through chronic, accelerated, and blast crisis phases, making disease stratification and therapeutic response prediction challenging. IGF2BP3 (Insulin-like Growth Factor 2 mRNA Binding Protein 3) has emerged as a potential prognostic biomarker due to its involvement in RNA stability and oncogenic pathways.

**Methods:** This study employed a multi-platform approach, including immunohistochemistry (IHC), ELISA, qRT-PCR, and Western blotting, to evaluate IGF2BP3 expression in 121 CML patient samples across disease stages. Advanced artificial intelligence (ChatGPT 4.0) and statistical tools (R-Studio) were utilized to analyze correlations between IGF2BP3 levels, clinical parameters, and treatment responses.

**Results:** IGF2BP3 expression progressively increased from the chronic to the blast crisis phase, correlating with disease severity and resistance to therapy. IHC staining intensity and serum levels of IGF2BP3 were highest in the blast crisis phase, confirmed by qRT-PCR and Western blotting. AI-powered regression analysis revealed a strong correlation between IGF2BP3 levels, P210 translocation percentages, and blast counts. Non-responders to therapy exhibited significantly elevated IGF2BP3 levels, underscoring its potential as a marker for treatment resistance.

**Conclusions:** IGF2BP3 is a reliable biomarker for CML disease progression, therapeutic resistance, and patient stratification. Targeting IGF2BP3 may offer novel therapeutic opportunities, particularly in advanced disease stages. This study demonstrates the utility of integrating AI in biomarker research to enhance precision and actionable insights.

## Introduction

Chronic Myeloid Leukemia (CML) is a clonal myeloproliferative disorder characterized by the presence of the Philadelphia chromosome and the BCR-ABL fusion gene, which drives the constitutive activation of tyrosine kinase, leading to uncontrolled cell proliferation ^1^. Despite advancements in tyrosine kinase inhibitors (TKIs) that have transformed the prognosis of CML patients, resistance to therapy and disease progression remain significant challenges ^2^. CML can progress from the chronic phase (CP) to accelerated phase (AP) and eventually to blast crisis (BC), often accompanied by poor prognosis ^3^. One of the reasons for this poor prognosis is the absence of early predictive biomarkers that can detect disease progression and treatment resistance ^4^. IGF2BP3 (Insulin-like Growth Factor 2 mRNA Binding Protein 3), an oncofetal RNA-binding protein, has emerged as a potential biomarker for aggressive cancers, including leukemia ^5,6^. The urgency of this research is underscored by the need for early predictive biomarkers to improve the prognosis of CML patients ^7^. IGF2BP3 plays a key role in regulating the stability and translation of mRNAs involved in cell growth and survival ^8^. Recent studies have shown that IGF2BP3 overexpression is correlated with poor survival and accelerated disease progression in several cancers, including leukemia ^9^. In particular, its overexpression is linked to increased proliferation, migration, and resistance to apoptosis in cancer cells ^10^. These findings suggest that IGF2BP3 could be a valuable target for therapeutic intervention and a potential prognostic marker for predicting treatment response and disease progression in CML. Despite its promising potential, there remains a need for more detailed studies investigating the specific role of IGF2BP3 in CML progression, particularly in its association with poor clinical outcomes. Identifying reliable molecular markers for CML progression and response to therapy is critical ^11^. This study aims to fill this gap by exploring the expression levels of IGF2BP3 in CML patients and evaluating its relationship with clinical outcomes, including survival and response to therapy ^12^. The potential impact of this study on patient care is significant, as it could lead to the development of early predictive biomarkers for CML progression and response to therapy, thereby improving patient outcomes ^13^.

In the era of precision medicine, advanced tools such as ChatGPT-4o ^14^ and R-Studio ^15^ offer novel approaches to analyzing complex datasets and predicting clinical outcomes ^16^. ChatGPT-4o, an advanced AI model, assists in generating predictive models to assess IGF2BP3’s role in disease progression. It analyzes large amounts of patient data to identify patterns and make predictions ^17^. Traditional bioinformatics tools such as R-Studio enable statistical data validation, ensuring that the results are reliable and accurate ^18^. By leveraging both AI-powered predictions and bioinformatics-driven analysis, this study provides comprehensive insights into the impact of IGF2BP3 on survival and disease progression in CML. Studies investigating the role of IGF2BP3 in CML are crucial for understanding how its overexpression contributes to treatment resistance and disease progression. Furthermore, this research is a collaborative effort, integrating AI-driven models from ChatGPT-4 and R-Studio’s statistical analysis, allowing for a comprehensive evaluation of IGF2BP3’s predictive value in CML progression and survival outcomes. This collaborative approach underscores the shared responsibility of the research community in advancing our understanding of CML and improving patient care.

How ChatGPT-4o Integrates into the Research:

- Predictive Modelling: ChatGPT-4 can generate predictive models (e.g., Random Forest, Gradient Boosting) based on patient data, identifying how IGF2BP3 overexpression influences survival rates and progression from chronic phase to blast crisis.
- AI-Driven Insights: By integrating multi-omics data, ChatGPT-4 can provide hypotheses regarding other potential biomarkers associated with IGF2BP3 expression, offering a deeper understanding of the molecular mechanisms at play in CML.
- Natural Language Summarization: ChatGPT-4 assists in generating readable, clinically relevant insights, helping clinicians and researchers understand complex bioinformatics findings related to IGF2BP3 and its impact on treatment outcomes.

Chronic Myeloid Leukemia (CML) research is advancing with a focus on identifying novel biomarkers like IGF2BP3, crucial for early detection and improved patient outcomes. IGF2BP3, known for stabilizing mRNAs linked to cell survival and growth, has shown strong associations with poor prognosis and cancer therapy resistance. Leveraging advanced tools like ChatGPT-4 and R-Studio enables comprehensive analysis of IGF2BP3’s role in CML progression. These technologies aid in predictive modeling, integrating multi-omics data, and uncovering actionable insights. By combining AI-driven predictions and bioinformatics validation, this research strives to enhance precision medicine, addressing critical gaps in understanding and managing CML progression.

## Methodology

### IGF2BP3 as a prognostic biomarker for cancer

We utilized multiple data sources and analytical tools to evaluate IGF2BP3 as a prognostic biomarker for cancer. The Smith and Sheltzer (2022) (https://www.tcga-survival.com/data-table?view=gene) research paper was referenced for z-score analysis to identify IGF2BP3’s association with poor prognosis. Differential gene expression analysis for IGF2BP3 was conducted using TIMER (Tumor IMmune Estimation Resource, available at https://cistrome.shinyapps.io/timer/). Additionally, the TCGA GDC portal (https://portal.gdc.cancer.gov/genes/ENSG00000188493) was used to extract information on IGF2BP3, specifically on gene alterations such as gain or loss of function. These resources comprehensively understood IGF2BP3’s role across various cancer types.

### Patient Samples and Processing

One hundred twenty-one (121) human CML samples and 30 healthy control samples were collected from the Departments of Pathology and Biochemistry at King George’s Medical University, following all participants’ ethical approval (*Reference Code-XVI-PGTC-IIA/P34*) and informed consent. The clinical diagnosis of CML was confirmed through patient history, peripheral blood smear examination, and bone marrow analysis using Leishman staining. Immunophenotyping of blast cells was conducted via flow cytometry using markers such as CD34, CD33, CD14, CD20, CD10, CD19, HLA-DR, TdT, CD2, CD3, CD5, CD7, CD13, CD23, CD45, CD64, CD79a, CD117, and CD200. Detailed patient characteristics are summarized in Table 1. All experiments were carried out promptly after sample collection in the Department of Biochemistry. Blood and bone marrow samples were aliquoted and preserved in a complete RPMI 1640 medium (Gibco) and stored in liquid nitrogen for long-term use. Before experiments, frozen cells were thawed and suspended in prewarmed RPMI 1640 supplemented with 40% FBS at 37°C. After thawing, the cells were washed and recovered for 45 minutes in the same medium at 37°C. Subsequently, the cells were rewashed and resuspended in PBS containing 0.1% BSA for downstream analyses.

**Table 1:** Clinical characteristics of CML patients.

| Characteristic of Patients | Examination Report |
| --- | --- |
| Normal | 30 |
| CML | 121 |
| **Age** |  |
| <40 | 81 (66.9) |
| >40 | 40 (33.1) |
| **Gender** |  |
| Male/Female | 75/46 |
| **Marital Status** |  |
| Married/Unmarried | 91/30 |
| **Average TLC** | Approx. 130000 |
| **Average Blast %** | Approx. 10% |
| **Spleen size** | Approx. 14 cm |
| **Phases** |  |
| Chronic Phase | 82 |
| Accelerated Phase | 19 |
| Blast Phase | 20 |
| **BCR ABL Translocation** |  |
| P210 Translocation | Approx. 40% |
| **Combination of marker** |  |
| **Treatment Response** |  |
| **Responsive** |  |
| Hematological basis | 31 |
| Molecular basis | 76 |
| **Non-Responsive** |  |
| Hematological basis | 11 |
| Molecular basis | 2 |

### Peripheral Blood Smear and Bone Marrow Staining

Leishman stain was prepared by dissolving 0.2 g of Leishman powder in 100 mL of methanol. After preparation, blood films were air-dried and subsequently stained with Leishman stain for 2 minutes. The slides were washed with buffered water for another 2 minutes to achieve a pinkish tinge. Blood films prepared from buffy coat samples were similarly stained for cases with suspected leukemia using Leishman stain for enhanced diagnostic accuracy ^19^.

### Confirmation of Blast Cells by Flow Cytometry

For flow cytometry analysis, monoclonal antibodies conjugated with fluorescein isothiocyanate (FITC), phycoerythrin (PE), peridinyl chlorophyllin (PerCP), and phycoerythrin-Cy7 (PE-Cy7) fluorochromes were used. Antibodies such as anti-CD45 V500-A, anti-CD34 PercP-A, anti-CD33 APC-A, anti-CD79a APC-A, anti-CD13-PE-A, anti-CD19-PE, anti-CD25-FITC-A, anti-CD20-V450-A, anti-CD7-FITC-A, anti-CD10-PE, anti-CD14-APC, anti-CD64 FITC-A, anti-CD117-PE, and HLA-DR-APC were procured from BD Biosciences (San Jose, CA, USA). The stain-lyse-wash method was employed for sample preparation. FACS tubes were labeled with patient identifiers and antibody combinations. For each tube, 100 μL of the sample (either bone marrow aspirate or whole blood) was mixed with 20 μL of antibody or antibody cocktail and incubated in the dark for 10–15 minutes. Following incubation, 2 mL of diluted FACS lysing solution was added to each tube. Samples were centrifuged at 200–300 g for 3–5 minutes, and the supernatant was discarded. The cell pellet was broken and washed twice with sheath fluid to remove residual debris. The final cell suspension was prepared by resuspending the cells in 0.5 mL of sheath fluid. Data acquisition was performed using a pre-calibrated FACSCanto flow cytometer (BD Biosciences), ensuring high-quality data collection and accurate blast cell characterization.

### Protocol for CytoCell^®^ BCR/ABL (ABL1) Translocation Detection Using FISH

The CytoCell^®^ BCR/ABL (ABL1) Translocation, Dual Fusion Probe is a fluorescence in situ hybridization (FISH) assay used to detect chromosomal rearrangements between 22q11.2 on chromosome 22 and 9q34.1 on chromosome 9. Bone marrow or peripheral blood samples were fixed in Carnoy’s solution (3:1 methanol/acetic acid) and prepared as monolayers on glass slides. Slides were pre-treated in 2X SSC buffer at 37°C for 10 minutes, followed by pepsin digestion at 37°C to enhance probe accessibility. Slides were air-dried after washing in 2X SSC and dehydration through an ethanol series. The BCR/ABL probe was denatured at 75°C, applied to the target area, and hybridized overnight at 37°C in a humidified chamber. Post-hybridization washes include 0.4X SSC/0.3% NP-40 at 72°C and 2X SSC/0.1% NP-40 at room temperature, followed by counterstaining with DAPI. Fluorescence microscopy analyzed fusion signals, with overlapping red and green signals indicating the BCR/ABL translocation. Positive and negative controls ensure accuracy. The assay supports diagnosing chronic myeloid leukemia (CML), acute myeloid leukemia (AML), and acute lymphoblastic leukemia (ALL), enabling precise detection of genetic abnormalities crucial for patient management. Biosafety precautions were observed throughout the process to ensure sample integrity and operator safety.

### Immunohistochemistry (IHC) Protocol

Tissue sections were fixed at 60°C on a hot plate for 60 minutes. The slides were then deparaffinized by immersion in xylene for 15 minutes, followed by xylene-alcohol treatment for 5 minutes. Rehydration was performed through a graded alcohol series: 100%, 90%, 70%, 50%, and 30% ethanol, each for 5 minutes, and slides were washed twice with distilled water (5 minutes each). Antigen retrieval was carried out in Tris-EDTA buffer (pH 9) at 110°C for 20 minutes, and the slides were cooled to room temperature. After this, the slides were washed twice with TBS buffer (pH 7.4) for 5 minutes.

To block endogenous peroxidase activity, the slides were incubated with peroxidase blocking solution for 15 minutes, followed by two washes with TBS buffer. Primary antibody incubation was performed for 1 hour at room temperature. After washing twice with TBS buffer, the slides were incubated with a secondary antibody. The signal was visualized using DAB (1:100 ratio) for 10 minutes, and the slides were rinsed with a TBS buffer. Hematoxylin staining was performed for 1–5 minutes, followed by washing under running water. Dehydration was carried out through graded alcohol series (70%, 90%, and 100%), followed by clearing in xylene. Finally, the slides were mounted for microscopic evaluation.

### Immunohistochemistry Interpretation

The staining intensity was graded as follows: non-existent (0), weak (1), moderate (2), and strong (3). The percentage of stained cells was scored as follows: no cell stained (0), <10% (1), 10–50% (2), 50–80% (3), and >80% (4). The final score was calculated by multiplying the staining intensity and the percentage of cells stained. A total score of 0–5 was interpreted as negative (IGF2 not overexpressed), while a score ≥6 was considered positive (IGF2 overexpression).

### Enzyme-Linked Immunosorbent Assay (ELISA)

Serum was collected from whole blood samples in plain vials and analyzed using commercially available ELISA kits (Human Insulin-Like Growth Factor 2 Mrna-Binding Protein 3 (Igf2Bp3) Elisa Kit from Biomatik (EKC41652)). All assays were performed according to the manufacturer’s instructions. The optical density was measured using a microplate reader, and the concentrations of target proteins were calculated based on standard curves provided with each kit.

### Gene Expression Analysis

Total RNA was isolated from cells using the TRIzol method, and RNA concentration and integrity were assessed with a Nanodrop 2000 UV–Vis spectrophotometer (Thermo Scientific). Only RNA samples with a 260/280 nm absorbance ratio of 1.9–2.0 were used. The extracted mRNA was reverse-transcribed into cDNA using the High-Capacity Reverse Transcription Kit (4368814) per the manufacturer’s instructions. Quantitative reverse transcription-PCR (qRT-PCR) was conducted using PowerUp SYBR Green Master Mix (ABI-A25741) on a 7500 Fast Real-Time PCR System (Applied Biosystems, Thermo Scientific). Gene expression was quantified using the ΔΔCt method, with β-actin as the reference gene. Data analysis was performed using DataAssist software (Thermo Scientific). Primer sequences are used for this IGF2BP3 F: CCTGGTGAAGACGGGCTAC; R: TCAACTTCCATCGGTTTCCCA; Here are commonly used β-actin primer sequences for qRT-PCR: Forward Primer: 5’-CATGTACGTTGCTATCCAGGC-3’ Reverse Primer: 5’-CTCCTTAATGTCACGCACGAT-3’ The specificity of the primers was confirmed by melting curve analysis and 2.2% agarose gel electrophoresis. The qRT-PCR protocol consisted of 40 cycles: 15 seconds at 95°C, 15 seconds at the annealing temperature (60°C for most genes), and 15 seconds at 72°C. Each experiment was performed in duplicate, with three independent biological replicates.

### Western Blotting

Proteins from blood and bone marrow samples were extracted using RIPA lysis buffer, and their concentrations were measured with the BCA protein assay at 562 nm using a spectrophotometer (Thermo Scientific). Protein samples were resolved on 15% SDS-polyacrylamide gels and transferred onto PVDF membranes (Bio-Rad). Membranes were incubated overnight at 4°C with primary antibodies, followed by washing and a 1-hour incubation with HRP-conjugated secondary antibodies. Immunoreactive protein bands were detected using the Odyssey Scanning System (LI-COR Inc., Lincoln, NE, USA). All experiments were repeated at least three times for reproducibility.

### Methodology for AI-Enhanced Analysis of IGF2BP3 Expression and Clinical Parameters in CML

This study utilized ChatGPT 4.0 and R-Studio to analyze IGF2BP3 expression about clinical parameters in Chronic Myeloid Leukemia (CML) patients. ChatGPT 4.0, with its advanced natural language processing (NLP) and data analysis capabilities, was employed to uncover correlations and trends from complex datasets. Scatter plot analyses identified relationships between P210 translocation percentages and IGF2BP3 expression, with regression models demonstrating a positive correlation. Stratification of IGF2BP3 levels by translocation ranges further emphasized its role in advanced CML stages.

Predictive modeling integrated with ChatGPT analyzed the association of IGF2BP3 with bone marrow blast percentages, confirming its relevance as a marker for disease severity. Additionally, ChatGPT-powered trend analysis revealed stage-wise IGF2BP3 expression, correlating with disease progression from chronic phase to blast crisis. Classification analysis highlighted elevated IGF2BP3 levels in treatment non-responders, suggesting its utility in predicting therapy resistance.

R-Studio was used for statistical validation and visualization. Heatmaps and correlation analyses quantified relationships between IGF2BP3 expression, immunohistochemistry (IHC) scores, and other clinical parameters, with strong correlations observed for IHC intensity (R = 0.81). Quantitative data, presented as mean ± standard deviation, were derived from at least three biological replicates, ensuring reliability. This dual AI and statistical approach provided robust insights into IGF2BP3’s role as a prognostic biomarker in CML.

### Statistical Analysis

Statistical analyses were conducted using GraphPad Prism 9 and SPSS version 16.0. The methods included Student’s t-test for pairwise comparisons, one-way ANOVA for multiple group comparisons, Chi-square test for categorical variables, Pearson correlation for relationship analysis, and significance levels were denoted as follows: *p < 0.01, **p < 0.001, ***p < 0.0001, and ****p < 0.00001. Quantitative data are expressed as mean ± standard deviation (SD) and were derived from at least three independent samples per data point to ensure reliability and reproducibility.

## Results

### Identification of IGF2BP3 as a Prognostic Biomarker in Cancer

To identify key prognostic biomarkers across various cancers, we analyzed data from the research article by Smith and Sheltzer, which performed genome-wide identification of prognostic features using z-score distributions across genes. Among the analyzed genes, IGF2BP3 emerged as a top candidate associated with poor prognosis, as shown in Figure 1A and Table S2. Specifically, IGF2BP3 demonstrated the highest positive z-score, signifying its strong association with patient mortality. In contrast, genes with lower z-scores were associated with improved survival outcomes. Further evaluation of the result highlighted IGF2BP3’s consistent identification as an informative feature across multiple platforms, including transcriptomic and proteomic datasets. The histograms presented in Figures 1B and 1C illustrate the comparative contributions of these platforms to the prognostic significance of IGF2BP3 and the robust evidence linking IGF2BP3 to adverse outcomes strongly emphasizes its relevance as a candidate for further investigation. Consequently, we chose to explore the specific role of IGF2BP3 in Chronic Myeloid Leukemia (CML), focusing on its contribution to disease progression and therapeutic potential. Here establishing the validity of this analysis conducted two types of Cox analysis using the processed clinical and genomic data as shown in Table S3 and S4. Univariate models, in which individual genomic features were directly associated with patient outcome, whereas multivariate models in which patient age, sex, tumor stage were incorporated along with genomic data. For each Cox model, as already reported the *Z* score, which encodes both the directionality and significance of a survival relationship. *Z* scores across cancer types were combined using Stouffer’s method (Stouffer, 1949). A *Z* score >1.96 indicates that the presence or upregulation of a tumor feature is associated with shorter survival times at a p < 0.05 threshold, while a *Z* score < −1.96 indicates that the presence or upregulation of a feature is associated with longer survival times at a p < 0.05 threshold. In general, the *Z* scores produced by the univariate and fully adjusted models were highly concordant within individual data types (median R = 0.96) and individual cancer types (median R = 0.95), suggesting that few prognostic markers were affected by the inclusion of additional clinical variables ^20^.

**Figure 1:**
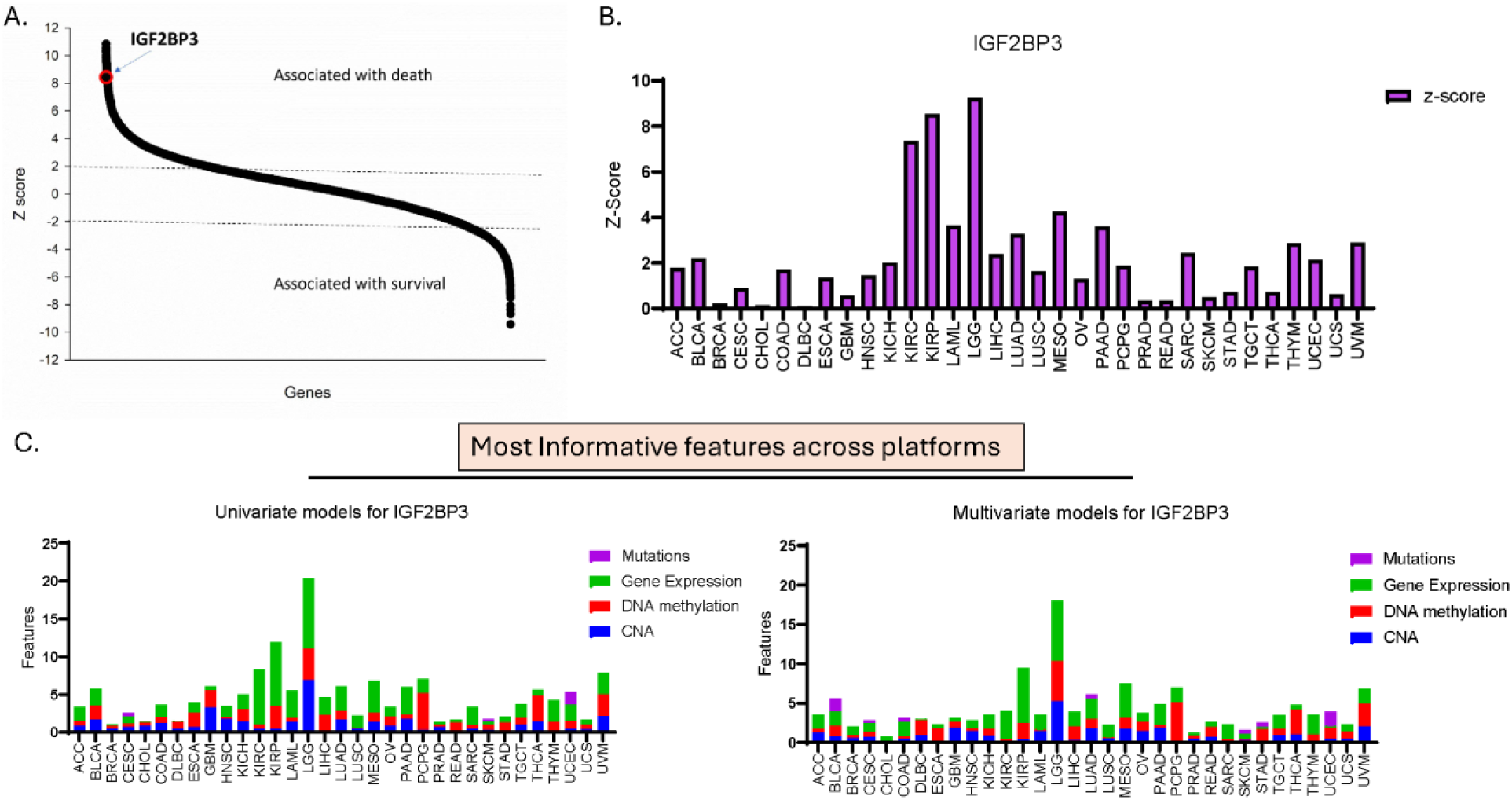
IGF2BP3 as a prognostic biomarker across cancers. **(A)** Z-score distribution for genes identified as prognostic biomarkers. IGF2BP3, with its highest z-score, signifies a robust association with patient mortality, underlining its crucial role in cancer progression. Genes with negative z-scores, on the other hand, are associated with better survival outcomes. **(B, C)** Contribution of IGF2BP3 to prognostic significance across various platforms. The bar plots depict the most informative features derived from transcriptomic, proteomic, and other datasets, with IGF2BP3 consistently demonstrating significance across these platforms.

### Differential Expression and Functional Analysis of IGF2BP3 in Various Cancer Types

The expression levels of IGF2BP3 were systematically analyzed across different cancer types, revealing significant upregulation in tumor tissues compared to their normal counterparts. As shown in Figure 2B, IGF2BP3 expression is markedly higher in several cancers, including BRCA, COAD, LIHC, LUAD, and PRAD, with statistical significance indicated (**p < 0.01, **p < 0.001). This consistent overexpression underscores IGF2BP3’s potential role in tumor progression and its utility as a diagnostic marker. The gain and loss of function of IGF2BP3 were further investigated to understand its functional implications in tumorigenesis. Figure 2B illustrates the percentage of cases with alterations, with a predominant gain of function observed in most cancer types. Further breakdown of these alterations is shown in the bottom panels: the gain-of-function events (2C) were more frequent across several cancer datasets, whereas the loss-of-function events (2D) were relatively less common, suggesting that gain-of-function mutations or overexpression may drive oncogenesis. Additionally, the summary of IGF2BP3’s genomic characteristics, including its synonyms and chromosomal location, is provided (Figure 2E) for reference. Together, these findings solidify IGF2BP3’s role as a cancer-associated gene with diagnostic and therapeutic relevance.

**Figure 2:**
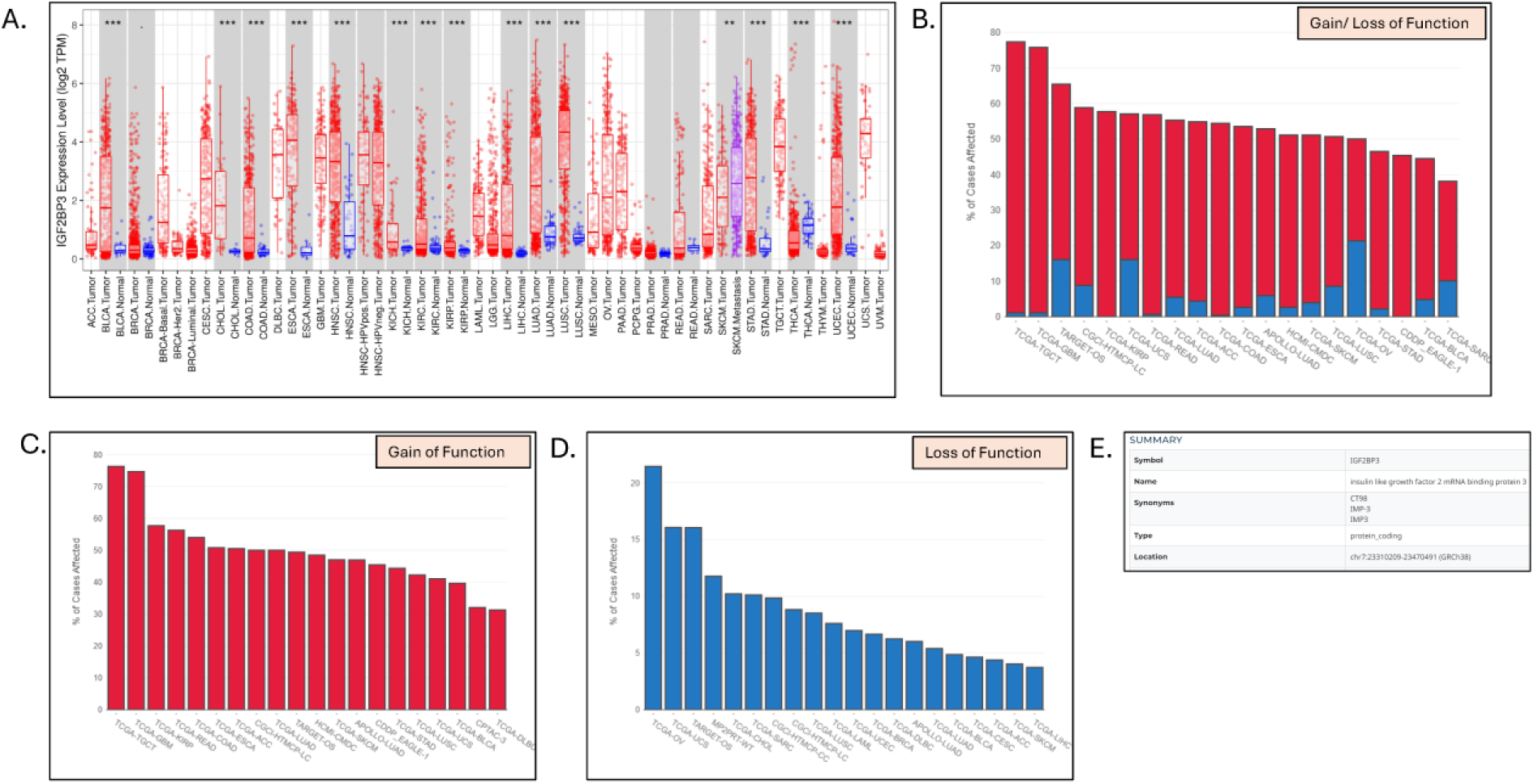
Differential expression and functional analysis of IGF2BP3 across cancers. **(A)** Boxplots representing IGF2BP3 expression (log2 TPM) in various tumor and routine tissue datasets. Significant overexpression of IGF2BP3 was observed in multiple cancer types. **(B)** The proportion of cases with gain- or loss-of-function alterations across cancer types. Red bars indicate gain-of-function events, while blue bars represent loss-of-function events. **(C)** Breakdown of gain-of-function events across datasets, with TCGA-LGG exhibiting the highest percentage. **(D)** Distribution of loss-of-function events across datasets shows lower frequencies than gain-of-function events. **(E)** IGF2BP3’s genomic details include synonyms, type, and chromosomal location (GRCh38).

### Morphological and Molecular Characterization of CML Blast Crisis

The clinical characteristics of 121 CML patients are summarized in Table 1 and Table S1. The cohort consisted of 75 males and 46 females, with a median age of 40 years. Most patients were in the chronic phase (67.8%), while a smaller fraction was in the accelerated (15.7%) or blast phase (16.5%). Hematological and molecular responses to treatment were observed in 31 and 76 patients, respectively, with non-responsiveness recorded in a minority. In this study, we analyzed the clinical and molecular characteristics of Chronic Myeloid Leukemia (CML) patients, including blood and bone marrow samples. The morphological evaluation of blast crisis cells from peripheral blood smears, bone marrow aspirates, and trephine biopsy specimens was performed, as shown in Figure 3A. The smears were stained using Leishman dye and revealed an abundance of myeloid blast crisis cells. Whereas Fluorescence in situ hybridization (FISH) analysis confirmed the presence of BCR-ABL translocation, as depicted in Figure 3B. This hallmark cytogenetic feature was observed in approximately 40% of the patients, supporting the diagnosis of CML in blast crisis. Flow cytometric immunophenotyping was performed on blast cells gated on CD45 vs. side scatter to evaluate the expression of various markers. The analysis revealed the presence of myeloid markers (CD13, CD33, CD117, CD14, CD64, cMPO), B-lymphoid markers (CD19, CD20, CD79a, CD10), T-lymphoid markers (CD3, cCD3, CD2, CD5, CD7), and immaturity markers (CD34, TdT, HLA-DR). Representative results are shown in Figure 3C, highlighting the phenotypic heterogeneity in CML patients. Approximately 95% of patients exhibited ∼15,000 blast cells out of 20,000 CD45 events, confirming the presence of blast cells.

**Figure 3:**
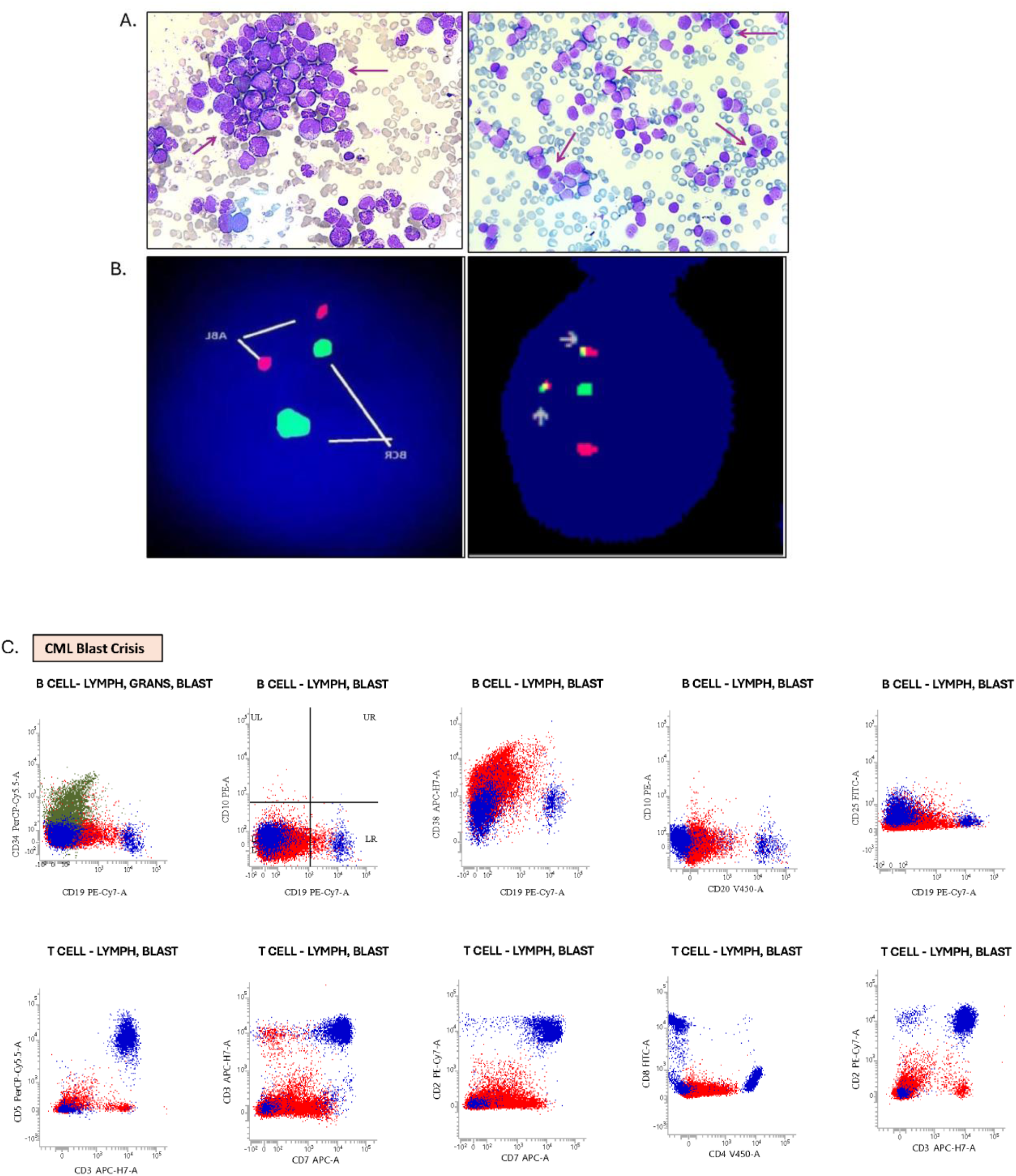
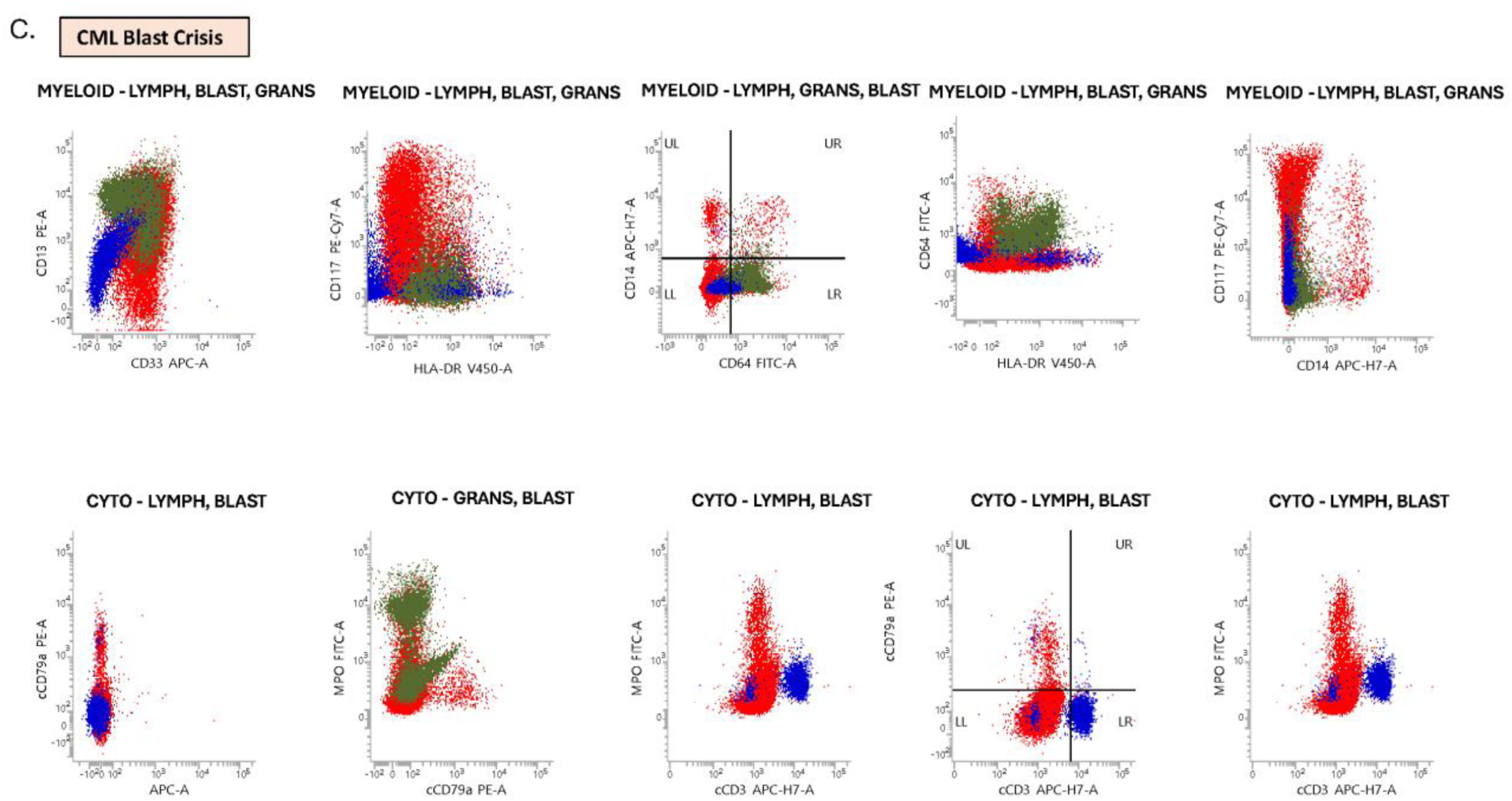
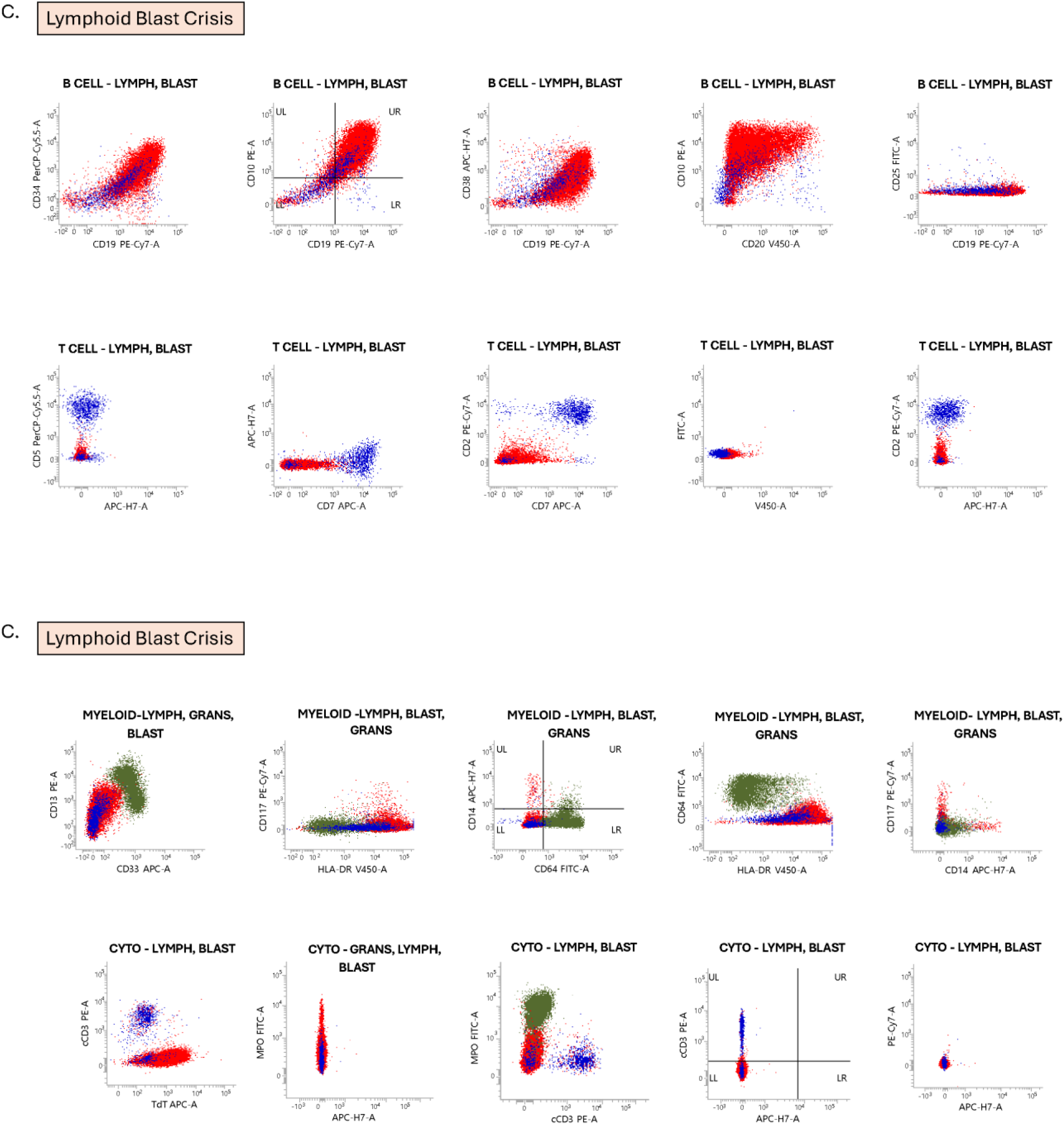
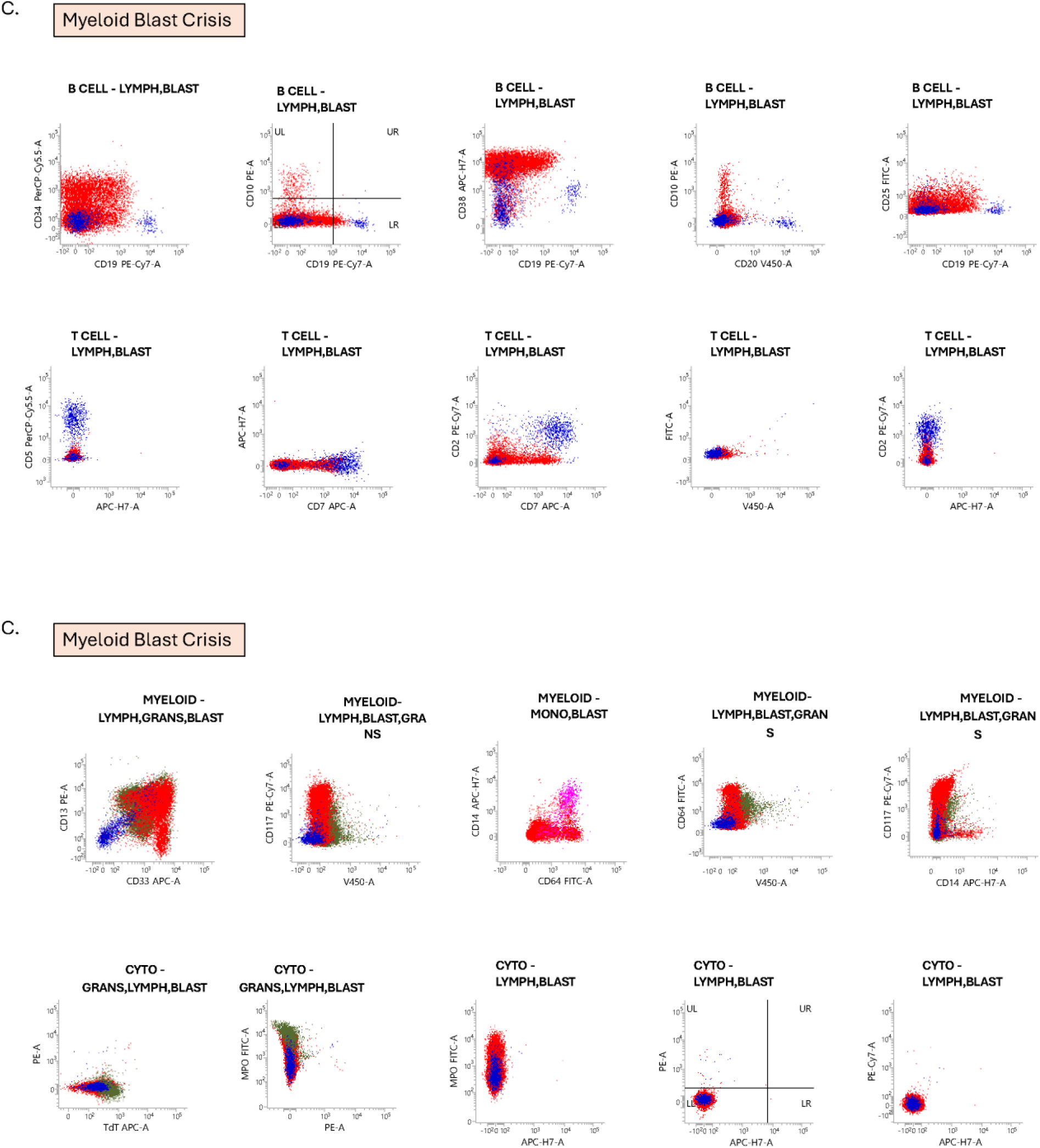
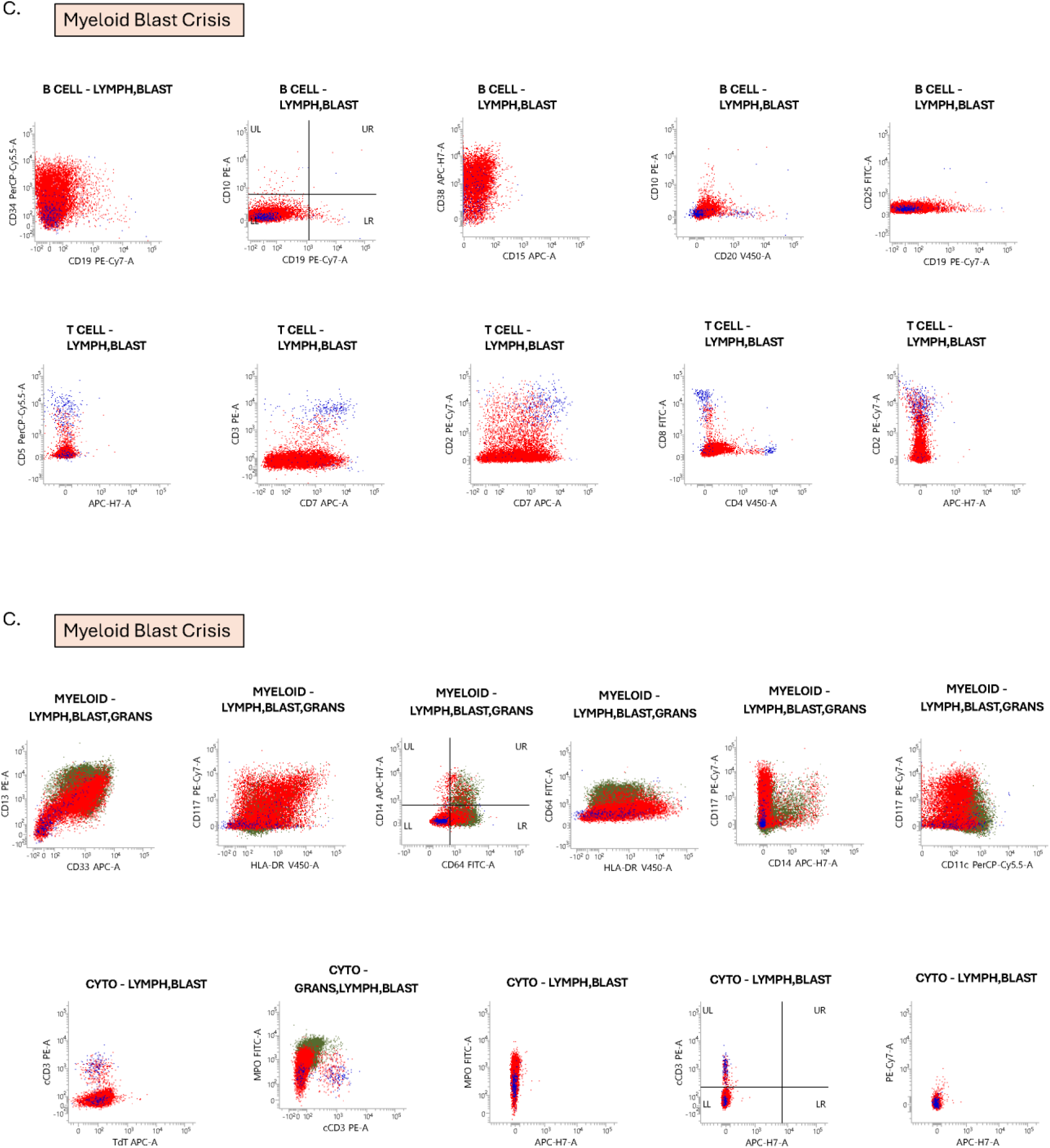
Morphological, molecular, and immunophenotypic characterization of CML blast crisis cells. **(A)** Peripheral blood smear and bone marrow aspirate smears stained with Leishman dye showing myeloid blast crisis cells (indicated by arrows). **(B)** Fluorescence in situ hybridization (FISH) analysis confirms the presence of BCR-ABL translocation (arrows point to the fusion signals). **(C)** Flow cytometry plots showing the expression of various myeloid, lymphoid, and immaturity markers in blast crisis cells. Different subsets, including B, T, and myeloid cells, are gated and analyzed for phenotypic heterogeneity.

### Stage-Wise Upregulation of IGF2BP3: A Biomarker of Disease Progression in Chronic Myeloid Leukemia

The analysis of IGF2BP3 expression across different stages of Chronic Myeloid Leukemia (CML)—chronic phase (CP), accelerated phase (AP), and blast crisis phase (BCP)—along with normal controls, revealed critical insights into the role of this biomarker in disease progression. Using multiple methodologies, including immunohistochemistry (IHC), ELISA, qRT-PCR, and Western blotting, the results demonstrated a consistent and progressive increase in IGF2BP3 expression with advancing CML stages.

IHC analysis provided a clear visual representation of IGF2BP3 expression at the tissue level. The staining intensity was weakest in the chronic phase (CP), moderate in the accelerated phase (AP), and strongest in the blast crisis phase (BCP), indicating a stage-wise upregulation of IGF2BP3. The placenta tissue, used as a positive control, demonstrated intense staining (+++), confirming the reliability of the assay. Quantitative assessment of IHC data revealed a steady increase in IGF2BP3 expression from CP to AP and peaking in BCP, suggesting its association with disease severity. These findings highlight the critical involvement of IGF2BP3 in the pathophysiological progression of CML as shown in Figure 4A. Protein-level analysis using ELISA further supported these observations. IGF2BP3 levels in serum were significantly higher in CML patients compared to healthy controls (p < 0.0001), with the most dramatic increase observed in the blast crisis phase. The progressive elevation of IGF2BP3 protein levels through the CP, AP, and BCP stages reinforces the hypothesis that IGF2BP3 expression correlates directly with disease advancement. This result was further validated by estimation plots, which confirmed significant differences in IGF2BP3 expression between normal and diseased samples as shown in Figure 4B. At the transcriptional level, qRT-PCR analysis revealed a robust upregulation of IGF2BP3 mRNA in CML patients compared to healthy controls. This upregulation of mRNA aligns well with the protein expression patterns observed through ELISA and IHC, suggesting that the enhanced transcription of IGF2BP3 could be driving its increased protein levels in advanced disease stages as shown in Figure 4C. Western blotting provided further validation at the protein level. The quantification of band intensities mirrored the trends observed in IHC, ELISA, and qRT-PCR, demonstrating the reliability and reproducibility of the findings across different analytical techniques as shown in Figure 4D. When the results from all methodologies were combined and compared, a consistent pattern emerged: IGF2BP3 expression was progressively upregulated from CP to AP, with the highest levels in BCP. This stage-dependent increase suggests that IGF2BP3 plays a crucial role in the transformation of CML from a relatively indolent state to an aggressive blast crisis phase. The findings indicate that IGF2BP3 could serve as a key biomarker for monitoring disease progression and as a potential therapeutic target.

**Figure 4:**
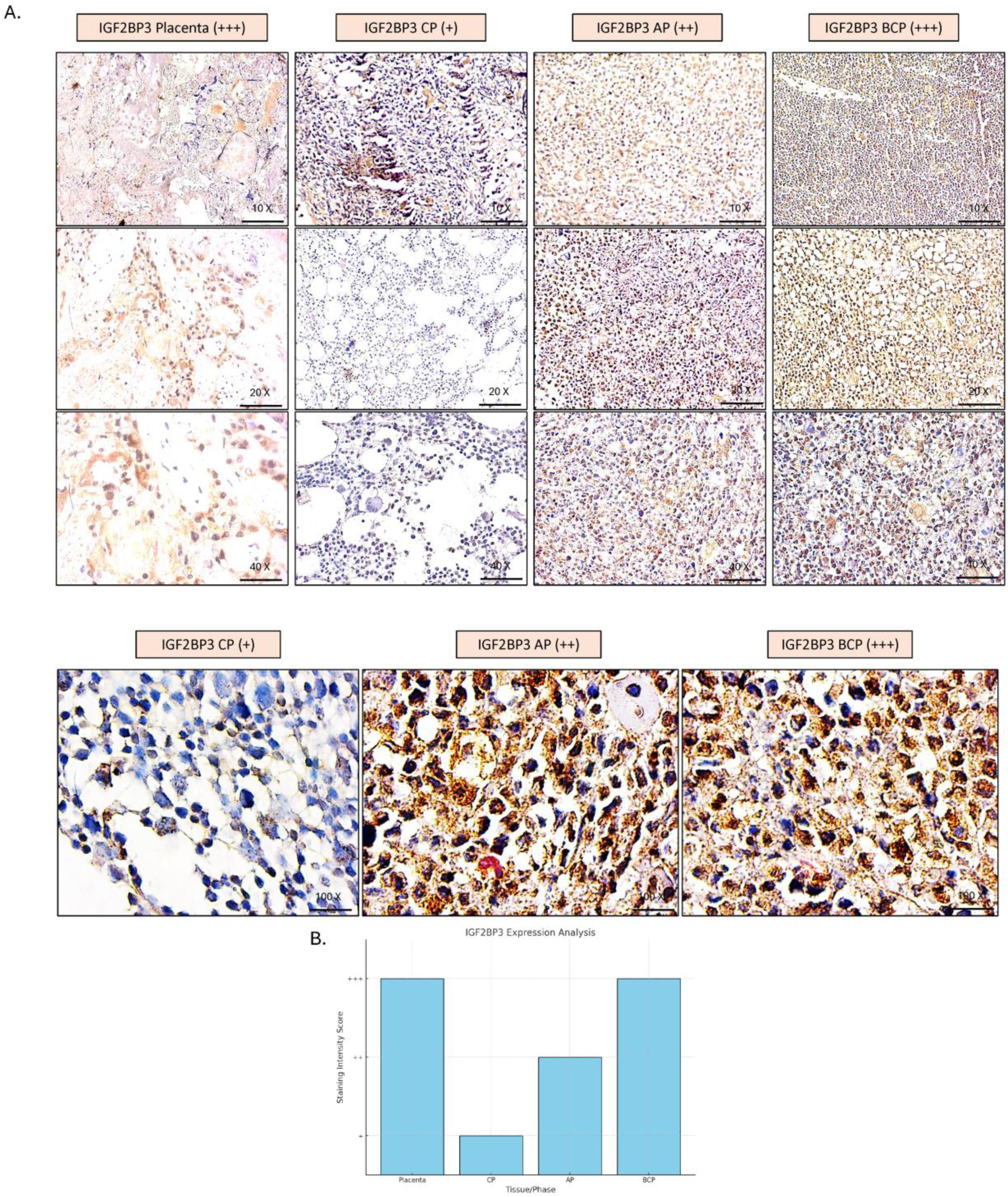
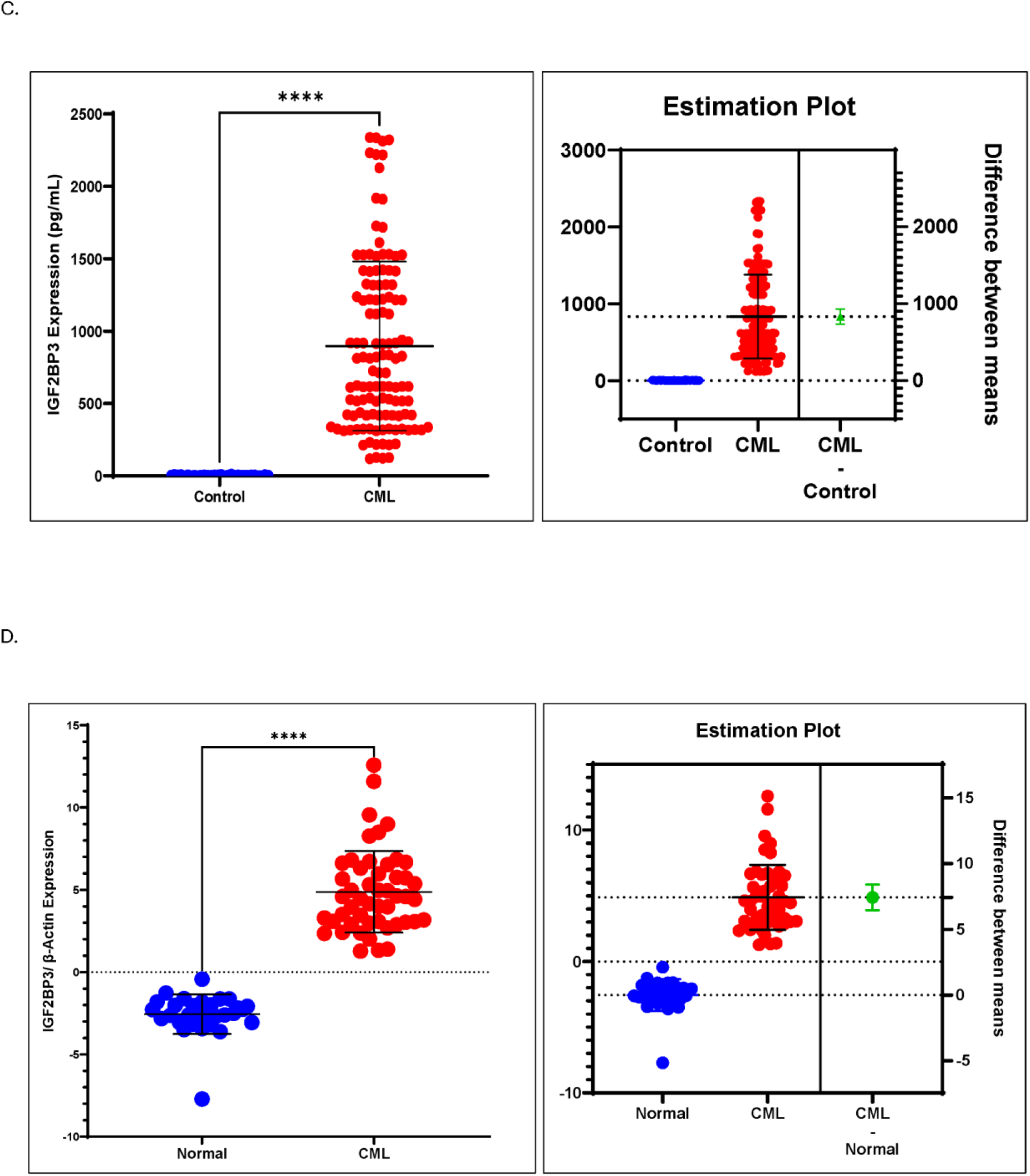
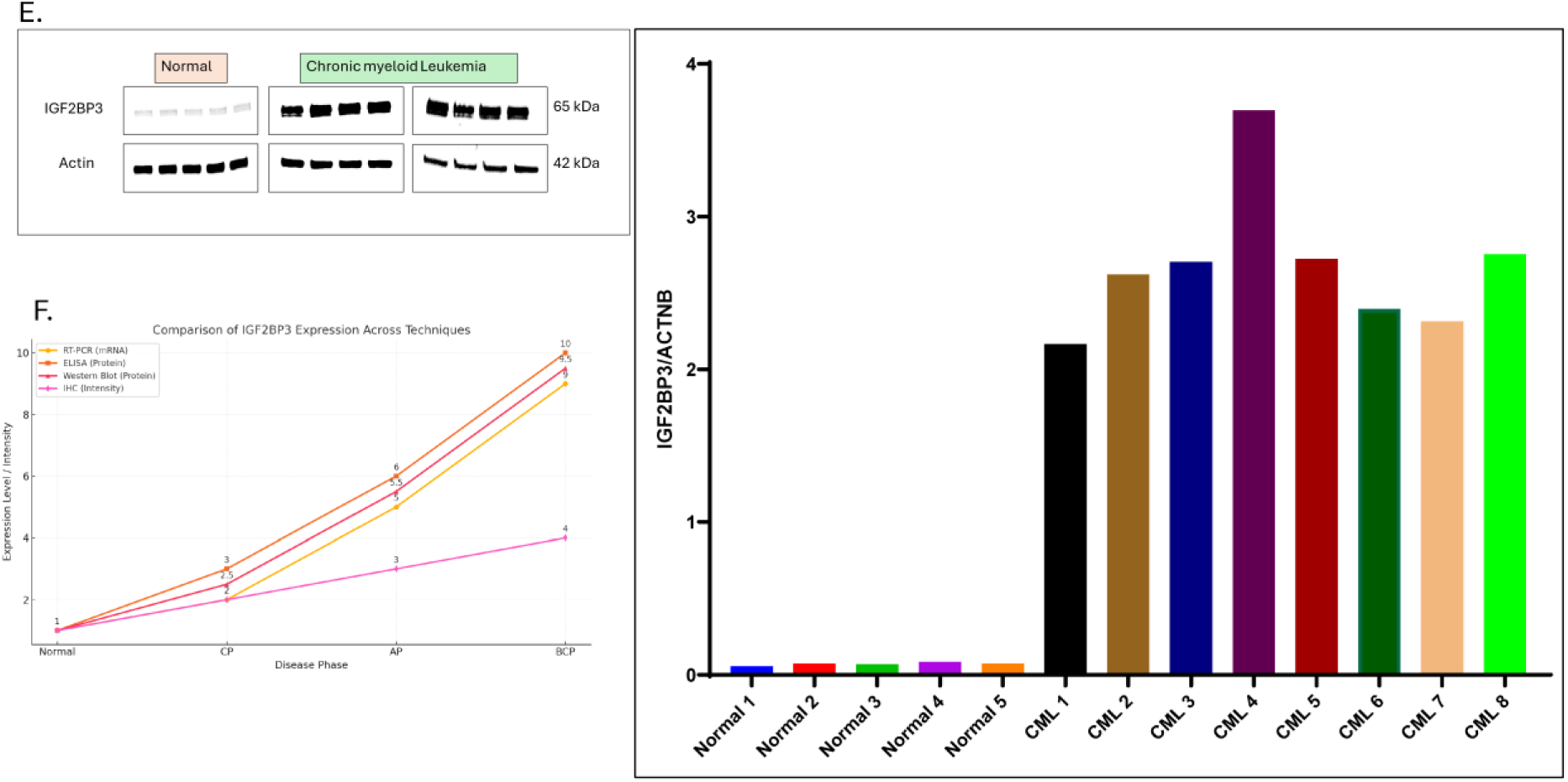
Differential expression of IGF2BP3. **(A)** Immunohistochemical (IHC) analysis of IGF2BP3 expression across different phases of CML. Representative images of IGF2BP3 staining in placenta (positive control), chronic phase (CP), accelerated phase (AP), and blast crisis phase (BCP) tissues. The progressive increase in staining intensity from CP to AP and its peak in BCP indicate a stage-wise upregulation of IGF2BP3, providing a clear pattern of disease progression. The images are shown at 10X, 20X, and 40X magnifications. **(B**) Quantitative analysis of IGF2BP3 IHC staining intensity. A bar graph shows the average staining intensity scores for IGF2BP3 in different phases of CML (CP, AP, and BCP) compared to placenta tissue (+++). The data confirm a progressive increase in IGF2BP3 expression with advancing disease stages. **(C**) ELISA analysis of IGF2BP3 protein levels in serum. Dot and estimation plots illustrate IGF2BP3 protein concentrations in serum samples from healthy controls and CML patients. A significant increase in IGF2BP3 levels is observed in CML patients, with the highest expression seen in the blast crisis phase (****p < 0.0001). **(D**) Quantitative real-time PCR (qRT-PCR) analysis of IGF2BP3 mRNA levels. Dot plot and estimation plot showing IGF2BP3 mRNA expression normalized to β-actin in routine controls and CML patients. A significant upregulation of IGF2BP3 mRNA is observed in CML, with levels increasing progressively from CP to AP and peaking in BCP (****p < 0.0001). **(E**) Western blot analysis of IGF2BP3 protein expression in CML. Western blot images showing IGF2BP3 expression in routine controls and CML samples across different disease stages (CP, AP, and BCP). β-actin was used as a loading control. The bar graph quantifies band intensities, demonstrating a progressive increase in IGF2BP3 expression with advancing disease stages. **(F**) Comparison of IGF2BP3 expression across different analytical techniques. Line graph comparing IGF2BP3 expression levels across normal controls and CML stages (CP, AP, and BCP) using RT-PCR, ELISA, Western blot, and IHC. All techniques consistently show a progressive upregulation of IGF2BP3 from normal to blast crisis phase, confirming its role in disease progression.

The result inferred into different implications; Biological Significance: The progressive increase in IGF2BP3 expression across CML stages suggests that it may play an active role in driving disease progression. The sharp upregulation in the blast crisis phase points to its involvement in the aggressive and proliferative characteristics of this stage. Diagnostic Utility: The consistent overexpression of IGF2BP3 in CML compared to normal controls, as demonstrated across multiple techniques, establishes it as a reliable biomarker for diagnosing and stratifying CML patients based on disease severity. Prognostic Potential: The correlation between IGF2BP3 levels and disease stages indicates its potential use as a prognostic marker. Patients with higher IGF2BP3 levels, particularly those in the accelerated or blast crisis phases, may have a poorer prognosis. Therapeutic Implications: The significant elevation of IGF2BP3 in blast crisis patients suggests that targeting this protein could be an effective strategy for controlling disease progression, particularly in advanced CML stages.

In conclusion, these results underscore the critical role of IGF2BP3 in the pathogenesis of CML and highlight its potential as a diagnostic, prognostic, and therapeutic target. Future studies focusing on the mechanistic role of IGF2BP3 and its interaction with key signaling pathways could provide deeper insights into its utility in CML management.

### ChatGPT 4.0-Enhanced Analysis of IGF2BP3 Expression with Clinical Parameters in CML Patients

To explore the relationship between IGF2BP3 expression and critical clinical parameters in Chronic Myeloid Leukemia (CML), ChatGPT 4.0 was employed to facilitate data interpretation and generate actionable insights. By leveraging its advanced natural language processing (NLP) and data analysis capabilities, ChatGPT enabled the identification of significant correlations and patterns from complex datasets that would be challenging to discern using traditional methods.

The scatter plot in Figure 5B highlights a positive correlation between P210 translocation percentage and IGF2BP3 levels. This relationship, identified through ChatGPT-enhanced regression analysis, was further quantified to show that increasing P210 translocation percentages correspond to elevated IGF2BP3 expression. This finding suggests a genetic linkage between P210 translocation and IGF2BP3 upregulation, underlying CML progression. Additionally, Figure 5C illustrates IGF2BP3 levels across stratified P210 translocation ranges, as determined using ChatGPT-assisted clustering techniques. Higher translocation percentages (60–100%) were associated with significantly elevated IGF2BP3 levels, underscoring its role in advanced disease stages. Whereas, Figure 5E demonstrates the relationship between bone marrow blast percentages and IGF2BP3 expression. Using predictive modeling integrated with ChatGPT, the analysis showed that higher blast percentages are strongly associated with increased IGF2BP3 levels. This emphasizes IGF2BP3 as a marker of disease severity, particularly in patients exhibiting aggressive phenotypes. ChatGPT’s capability to process and integrate diverse data points ensured accurate correlation and interpretation. While Figure 5F shows, the progressive increase in IGF2BP3 expression across different CML phases (chronic, accelerated, and blast crisis) was identified through ChatGPT-powered trend analysis. This finding was further validated through ChatGPT-aided quantification of IHC scores (Figure 5G), which revealed increased staining intensity correlating with disease progression. These AI-powered insights ensure precision, reproducibility, and objectivity in evaluating IGF2BP3 as a biomarker for tracking CML stages.

**Figure 5:**
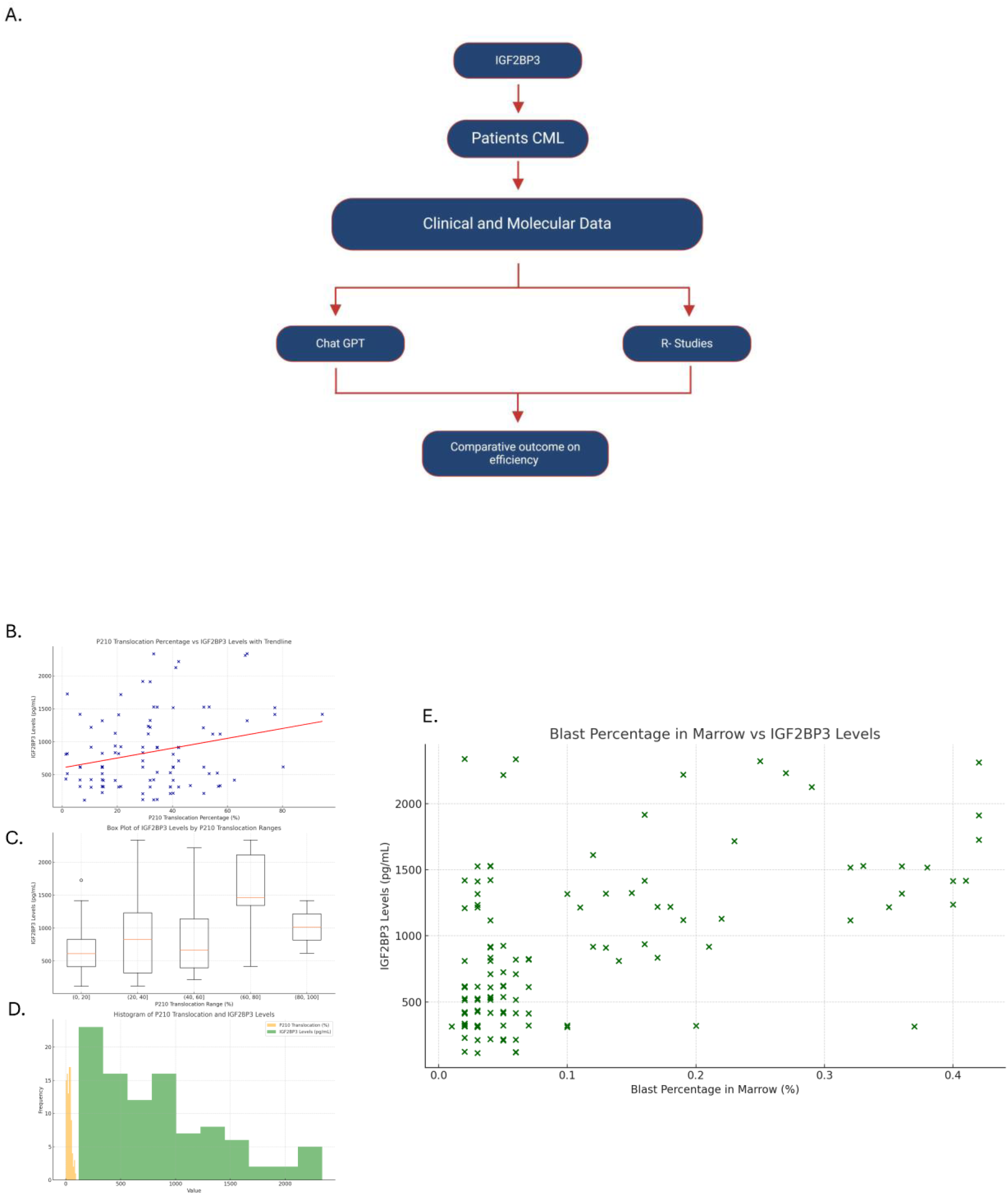
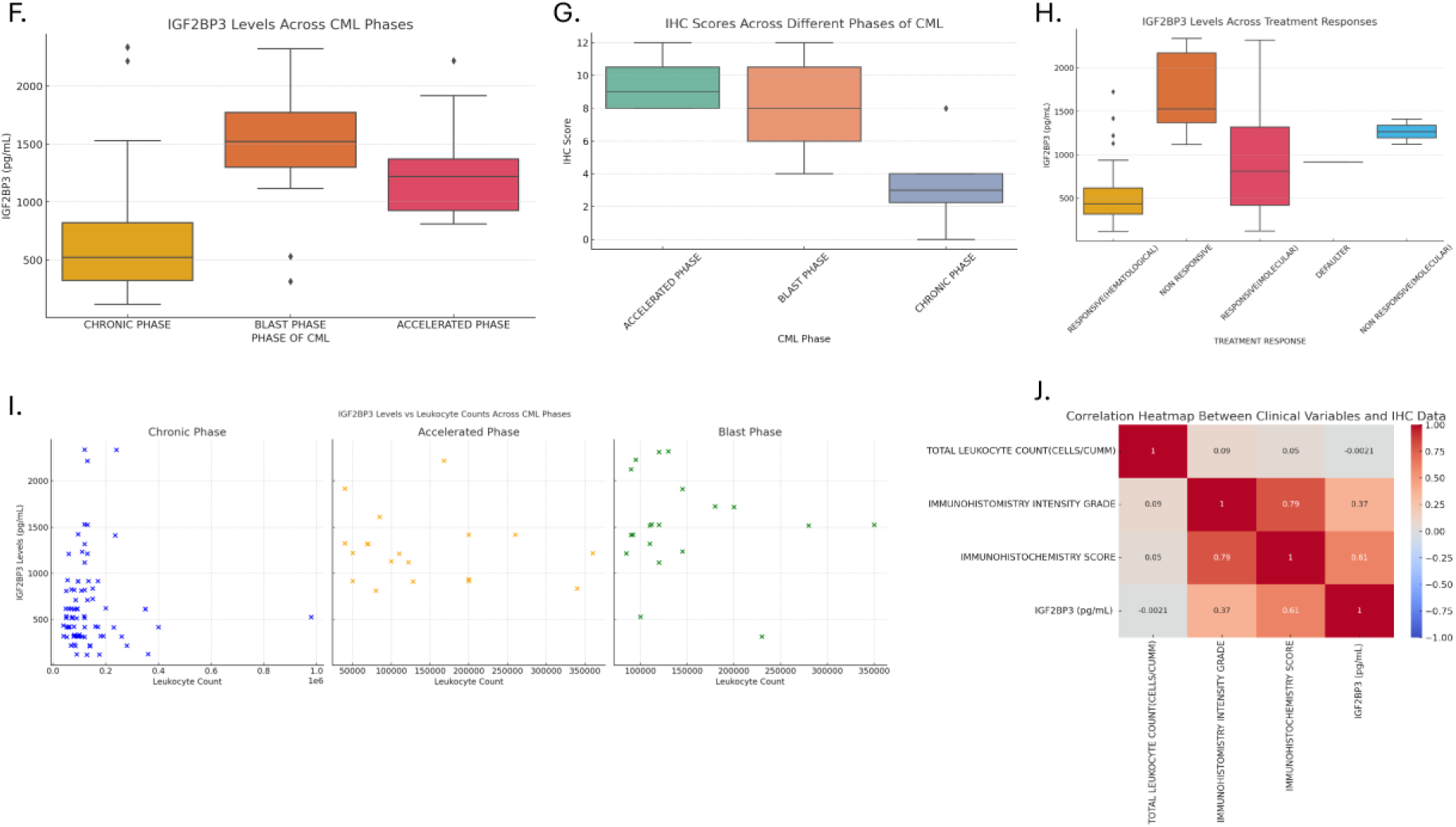
Comprehensive Analysis of IGF2BP3 Expression and Its Correlation with Clinical and Molecular Parameters in Chronic Myeloid Leukemia (CML). **(A)** Workflow depicting the analysis pipeline for IGF2BP3 in Chronic Myeloid Leukemia (CML). Clinical and molecular data from CML patients were analyzed using ChatGPT-based AI tools and R-Studies for comparison, culminating in insights into their efficiency in identifying significant patterns. **(B)** Correlation plot showing the relationship between P210 translocation percentage and IGF2BP3 levels (pg/mL). A positive trendline highlights a strong correlation, indicating that higher P210 translocation is associated with elevated IGF2BP3 levels. **(C)** Box plot representing IGF2BP3 levels across different P210 translocation ranges. Median IGF2BP3 levels increase with higher translocation ranges, emphasizing its potential role in disease progression. **(D)** Histogram illustrating the frequency distribution of P210 translocation percentages and IGF2BP3 levels. The graph demonstrates a clear stratification, with high IGF2BP3 levels clustering in patients with advanced translocation percentages. **(E)** Scatter plot correlating IGF2BP3 levels (pg/mL) with blast percentage in the bone marrow. A significant increase in IGF2BP3 levels is observed as blast percentages rise, suggesting its association with advanced disease phases. **(F)** Box plot of IGF2BP3 levels across different CML phases (chronic, accelerated, and blast). The highest IGF2BP3 expression is observed in the blast phase, followed by the accelerated phase, highlighting its role as a marker of disease progression. **(G)** Immunohistochemistry (IHC) score distribution for IGF2BP3 across different CML phases. Elevated IHC scores in the accelerated and blast phases further validate IGF2BP3’s upregulation in advanced disease stages. **(H)** Box plot comparing IGF2BP3 levels among different treatment response groups. Non-responders exhibit significantly higher IGF2BP3 levels, underscoring its potential as a marker for therapeutic resistance. **(I)** Scatter plots showing the correlation between IGF2BP3 levels and leukocyte counts across CML phases. Elevated IGF2BP3 levels correlate with increasing leukocyte counts, particularly in the blast phase. **(J)** Heatmap illustrating the correlation between clinical variables (total leukocyte count, IHC intensity grade, IHC score, and IGF2BP3 levels). Strong positive correlations are observed between IGF2BP3 levels, IHC intensity, and IHC score, highlighting their interdependence.

Figure 5H depicts treatment response analysis, where ChatGPT-enhanced classification revealed significantly higher IGF2BP3 levels in non-responsive patients, particularly at the molecular level. This suggests that IGF2BP3 could serve as a predictive marker for treatment resistance, enabling more personalized treatment approaches.

ChatGPT was also utilized in multivariate analysis (Figure 5I) to examine the relationship between IGF2BP3 levels and leukocyte counts across CML phases. The analysis identified minimal correlation in the chronic phase but stronger associations in the accelerated and blast phases, where elevated IGF2BP3 levels paralleled higher leukocyte counts, indicative of disease aggressiveness.

Finally, the correlation heatmap in Figure 5J was generated using ChatGPT-driven data integration, consolidating relationships between IGF2BP3 expression, IHC scores, and other clinical parameters. The analysis revealed strong positive correlations between IGF2BP3 levels and IHC intensity grade (R = 0.79) and immunohistochemistry scores (R = 0.61), highlighting its central role as a biomarker. ChatGPT’s ability to synthesize and present these relationships with unparalleled clarity and depth underscores its potential as a tool for biomedical research. By employing ChatGPT 4.0 for data analysis, we refined our understanding of IGF2BP3 expression and its clinical implications in CML. The model enabled us to uncover novel patterns and correlations, providing a robust and reproducible framework for analyzing IGF2BP3’s diagnostic, prognostic, and therapeutic potential. This approach demonstrates the utility of AI in accelerating biomedical discoveries, enhancing precision, and fostering data-driven insights in clinical research. ChatGPT’s role in integrating complex datasets and facilitating predictive analysis offers promising avenues for advancing personalized medicine in CML management.

### R-Studio Analysis of IGF2BP3 with Clinical Parameters Correlation Analysis with Clinical Features

The correlation heatmaps in Figures 6A and 6B provide a comprehensive overview of the relationships between IGF2BP3 levels and clinical parameters such as blast percentage in marrow, leukocyte counts, spleen size, and treatment response (BCR-ABL transcript levels). The analysis revealed:

- A strong positive correlation between IGF2BP3 levels and immunohistochemistry (IHC) scores (R = 0.81) and IHC intensity grade (R = 0.77), emphasizing the reliability of IGF2BP3 as a biomarker detectable through IHC.
- Moderate correlations with BCR-ABL transcript levels in non-responders (R = 0.66) and blast percentages in marrow (R = 0.55), suggesting a link between IGF2BP3 expression and aggressive disease markers.
- A weak inverse correlation with leukocyte count (R = −0.12), highlighting that IGF2BP3 levels are not solely dependent on leukocyte load but likely influenced by other disease mechanisms.

**Figure 6:**
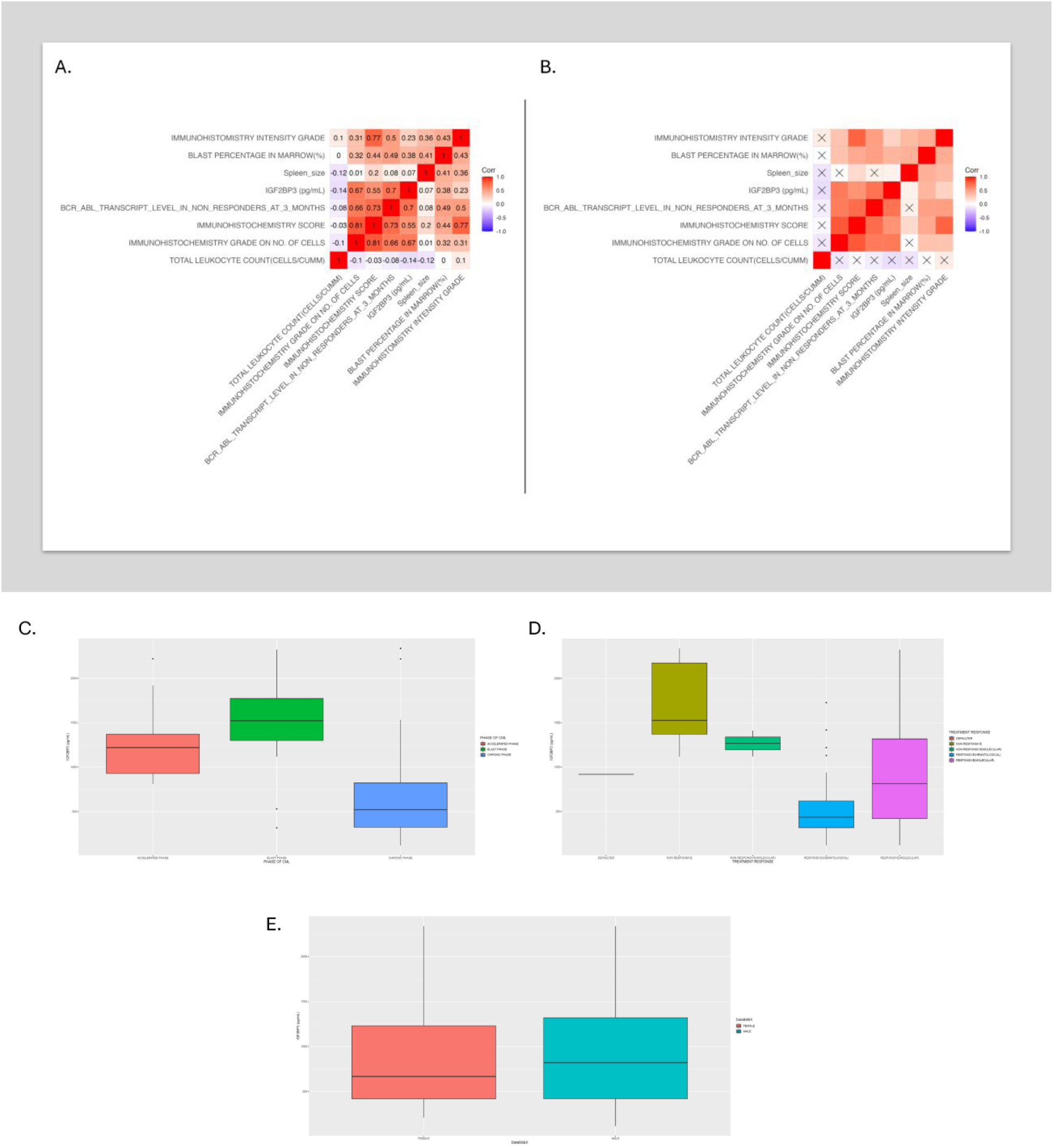
Correlation and Distribution Analyses of IGF2BP3 Levels in Chronic Myeloid Leukemia (CML). **(A)** Correlation heatmap depicting relationships between clinical and molecular parameters, including IGF2BP3 levels, total leukocyte counts, blast percentage in marrow, BCR-ABL transcript levels in non-responders, and immunohistochemistry (IHC) scores. Strong positive correlations are observed between IGF2BP3, IHC grades, and BCR-ABL levels. **(B)** The correlation heatmap with masked insignificant correlations highlights only statistically significant relationships among variables. In non-responders, IGF2BP3 levels strongly correlate with IHC intensity grades and BCR-ABL transcript levels. **(C)** Box plot showing IGF2BP3 levels across different CML phases (chronic, accelerated, and blast). The highest IGF2BP3 expression is observed in the blast phase, indicating its association with disease progression. **(D)** Box plot illustrating IGF2BP3 levels across different treatment response groups. Non-responders exhibit significantly higher IGF2BP3 levels than responders, emphasizing its potential role in therapy resistance. **(E)** Box plot comparing IGF2BP3 levels by sex (male and female). No significant difference indicates sex-independent expression of IGF2BP3 in CML patients.

The heatmaps provide a quantitative basis for interpreting the clinical significance of IGF2BP3, where its expression reflects both molecular changes and clinical manifestations in CML. Expression Trends Across Disease Phases In Figure 6C, IGF2BP3 levels are stratified across the chronic, accelerated, and blast phases of CML: A progressive increase in IGF2BP3 levels is observed from the chronic phase to the accelerated phase, with the highest expression in the blast phase. This trend is statistically significant, aligning IGF2BP3 with disease progression. The analysis highlights IGF2BP3 as a potential biomarker for monitoring disease stage and predicting transitions to more aggressive phases. Association with Treatment Response Figure 6D shows the distribution of IGF2BP3 levels based on patient response to treatment: Non-responsive patients exhibit markedly elevated IGF2BP3 levels compared to responders, particularly those responsive on a molecular basis. This trend suggests that IGF2BP3 may serve as a predictive marker for treatment resistance, aiding in stratifying patients for personalized therapeutic strategies. Sex-Based Differences in expression Figure 6E compares IGF2BP3 levels between male and female patients: While no significant sex-based differences were observed, the data reinforces the consistency of IGF2BP3 expression across demographics, enhancing its utility as a universal biomarker. The integration of R-Studio analysis enabled precise quantification and visualization of IGF2BP3 expression across various clinical parameters. ChatGPT-supported interpretation enriched the understanding of these correlations, providing actionable insights into IGF2BP3’s role as a prognostic marker in CML. This dual approach demonstrates the power of combining statistical rigor with AI-driven interpretation to generate robust, clinically relevant insights.

## Discussion

Chronic Myeloid Leukemia (CML) is a hematological malignancy characterized by its progressive phases: the chronic phase (CP), accelerated phase (AP), and blast crisis phase (BCP) ^20^. Despite significant advances in understanding the molecular pathogenesis of CML, early differentiation of disease stages and the prediction of patient prognosis remain critical challenges ^21^. The discovery and validation of robust biomarkers that can stratify patients across disease stages and predict therapeutic responses are essential for improving disease management ^22,23^. Among the myriad candidates explored in cancer biology, IGF2BP3 has emerged as a compelling target due to its consistent association with aggressive phenotypes in various malignancies ^6^. This study investigates the role of IGF2BP3 as a prognostic biomarker in CML, supported by advanced AI-model integration for precise and actionable insights. IGF2BP3 was selected based on its established role in oncogenesis and its strong association with poor prognosis in other cancers, as reported in genome-wide studies. Smith and Sheltzer’s analysis of prognostic biomarkers across 32 cancers identified IGF2BP3 as a top candidate with the highest z-score associated with mortality, underscoring its clinical relevance ^20^. This motivated us to explore IGF2BP3’s potential in CML, a disease that transitions from an indolent chronic phase to an aggressive blast crisis. Given the pressing need for biomarkers that can differentiate patients at various stages and predict outcomes, IGF2BP3 is an ideal candidate for investigation. IGF2BP3 is known to function as an RNA-binding protein that stabilizes oncogenic transcripts, promoting proliferation, metastasis, and chemoresistance ^5^. In the context of CML, where dysregulated signaling pathways and genomic instability drive disease progression, the overexpression of IGF2BP3 could play a pivotal role in shaping the aggressive phenotype of blast crisis cells. This hypothesis drove our comprehensive evaluation of IGF2BP3 expression across CML stages using a combination of immunohistochemistry (IHC), ELISA, qRT-PCR, and Western blotting. Our findings demonstrate a progressive increase in IGF2BP3 expression from CP to AP and peaking in BCP, consistent across all analytical techniques. The IHC analysis clearly represented this stage-wise upregulation, with staining intensity correlating strongly with disease severity. In CP, IGF2BP3 expression was relatively low, reflecting the indolent nature of this phase. In contrast, the accelerated phase exhibited moderate expression, signaling the onset of disease progression. The blast crisis phase showed the highest expression, aligning with the aggressive proliferation and differentiation blockade characteristic of this stage. The placenta tissue, used as a positive control, validated the assay’s reliability by consistently demonstrating intense IGF2BP3 staining. Protein-level analysis using ELISA further substantiated these findings, revealing significantly elevated IGF2BP3 levels in serum samples from CML patients compared to healthy controls. Notably, the highest protein levels were observed in BCP, reinforcing the association between IGF2BP3 expression and disease severity. Similarly, qRT-PCR analysis showed a gradual upregulation of IGF2BP3 mRNA across the three phases, with the highest levels in BCP. Western blotting corroborated these results, highlighting the reproducibility and robustness of our multi-platform approach. To enhance the precision of our analysis and uncover deeper correlations, we integrated advanced AI models into our workflow. ChatGPT 4.0, a state-of-the-art natural language processing and data interpretation tool, was employed to analyze complex datasets and generate actionable insights. This marks the first study to leverage AI for the comprehensive evaluation of IGF2BP3 in CML, setting a precedent for future research in biomarker discovery. The AI-enhanced regression analysis revealed a strong positive correlation between P210 translocation percentages and IGF2BP3 expression, suggesting a genetic linkage that drives disease progression. This correlation, validated through clustering techniques, showed that patients with higher P210 translocation percentages (60–100%) exhibited significantly elevated IGF2BP3 levels, emphasizing its role in advanced disease stages. Similarly, predictive modeling demonstrated that IGF2BP3 levels correlated strongly with bone marrow blast percentages, underscoring its utility as a marker of disease severity. AI-powered trend analysis highlighted the progressive increase in IGF2BP3 expression across CML stages, confirming its potential as a stage-specific biomarker. The integration of ChatGPT into IHC data quantification enabled precise and reproducible scoring of staining intensity, eliminating subjectivity and ensuring consistency. Furthermore, AI-driven classification identified higher IGF2BP3 levels in non-responsive patients, suggesting its potential as a predictive marker for treatment resistance. The correlation heatmap generated through AI-driven data integration consolidated relationships between IGF2BP3 expression and clinical parameters, such as leukocyte counts, blast percentages, and IHC scores. The strong positive correlations observed with IHC intensity grade and immunohistochemistry score further validated IGF2BP3 as a reliable biomarker detectable through standard pathological techniques. These AI-enabled insights enhanced the depth of our analysis and demonstrated AI’s transformative potential in biomedical research. The consistent upregulation of IGF2BP3 across CML stages underscores its biological significance in disease progression. As a prognostic biomarker, IGF2BP3 offers several advantages. Its strong association with disease severity makes it a valuable tool for stratifying patients and predicting outcomes. In clinical practice, measuring IGF2BP3 levels could enable early identification of patients at risk of progressing to the blast crisis phase, allowing for timely intervention. The therapeutic implications of IGF2BP3 are equally compelling. Its role in stabilizing oncogenic transcripts suggests that targeting IGF2BP3 could disrupt key pathways driving disease progression. Lin et al. demonstrated that IGF2BP3 knockdown sensitized leukemia cells to menin-MLL inhibitors, reducing leukemic initiating cells and enhancing differentiation ^24^ [24]. This provides a strong rationale for exploring IGF2BP3-targeted therapies in CML, particularly for patients with refractory or relapsed disease.

This study represents the first comprehensive analysis of IGF2BP3 in CML using a multi-platform approach integrated with AI-driven insights. Using IHC, ELISA, qRT-PCR, and Western blotting ensured the robustness and reproducibility of our findings, while AI models provided a new dimension of precision and depth. Incorporating ChatGPT 4.0 enabled us to uncover novel patterns and correlations, demonstrating the utility of AI in advancing biomarker research. The ability of IGF2BP3 to differentiate patients across CML stages and predict treatment resistance positions it as a unique and valuable biomarker. Its consistent upregulation in advanced disease stages highlights its potential as a therapeutic target, opening new avenues for personalized medicine in CML. While our findings establish IGF2BP3 as a promising biomarker, further research is needed to elucidate its mechanistic role in CML pathogenesis. Investigating its interactions with signaling pathways and its impact on the tumor microenvironment could provide deeper insights into its biological functions. Additionally, clinical trials evaluating IGF2BP3-targeted therapies are warranted to translate these findings into therapeutic applications. The integration of AI into biomarker research offers exciting possibilities for the future. By combining multi-omics data with AI-driven analysis, researchers can uncover hidden patterns and generate actionable insights with unprecedented precision as shown in the graphical abstract Figure 7. Expanding the use of AI in CML research could accelerate the discovery of novel biomarkers and therapeutic targets, ultimately improving patient outcomes.

**Figure 7:**
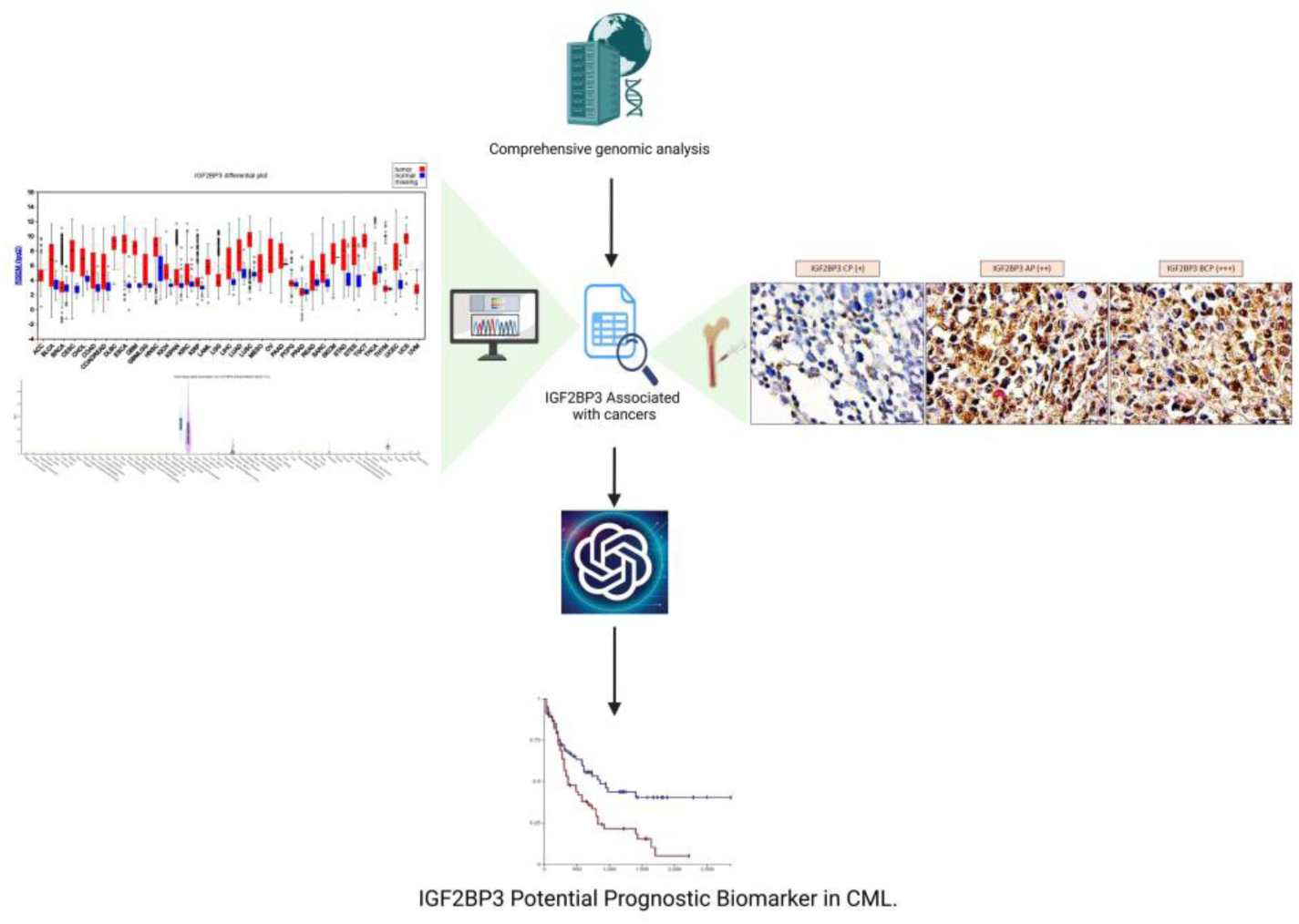
Graphical Abstract: IGF2BP3 has been identified as a critical prognostic biomarker in Chronic Myeloid Leukemia (CML) through comprehensive genomic analysis and immunohistochemical validation. Its elevated expression correlates with advanced disease stages and poor survival outcomes. Integrating advanced computational tools and survival data highlights its potential for targeted therapies and improved patient stratification.

## Conclusion

This study highlights IGF2BP3 as a critical biomarker in CML, with its expression correlating strongly with disease severity and treatment resistance. The progressive upregulation of IGF2BP3 across CML stages suggests its pivotal role in disease pathogenesis. AI-driven analysis further validated its potential as a prognostic and therapeutic target. These findings lay the groundwork for future research on IGF2BP3-targeted therapies and personalized treatment strategies in CML management.

## Data Availability

All data produced in the present work are contained in the manuscript

## Acknowledgments

This work was supported by the Department of Biotechnology, India (Grant No. BT/IN/Indo-US/Foldscope/39/2015). Dr. Vivek Singh (Principal Investigator) received funding for this research. The authors would like to thank Dr. Gauri Prasad (NCI-NIH USA) for performing data analysis using R-Studio and providing valuable insights for comparison. The authors also thank ChatGPT for assisting with analysis and enhancing the manuscript’s clarity.

## Contributions

VS and RK conceptualized and designed the experiments. PC, SU, and MK conducted the bench experiments and performed bioinformatics analyses. TS, GS, SPV, and RS organized the data and facilitated clinical validation. VS and RK wrote the manuscript.

## Ethics declarations

**Ethical approval**

**Institutional Ethics Approval Statement**

The research proposal entitled “To Decipher the Molecular Role of Insulin-like Growth Factor in Chronic Myeloid Leukemia” has undergone a thorough ethical review by the Institutional Ethics Committee (IEC) at King George’s Medical University, U.P., Lucknow (Registration No.: ECR/262/Inst/UP/2013/RR-19). The clarification provided for the comments raised during the initial review was evaluated, and the proposal has been approved for execution as per ethical guidelines. The decision reflects the compliance of the research objectives and methodology with established ethical standards for biomedical research. Researchers are requested to cite the assigned reference code: XVI-PGTSC-IIA/P34 in all future correspondence related to this study. For further queries or communication, the ethics committee can be contacted at King George’s Medical University, U.P., Lucknow.

